# Development of meta-prompts for Large Language Models to screen titles and abstracts for diagnostic test accuracy reviews

**DOI:** 10.1101/2023.10.31.23297818

**Authors:** Yuki Kataoka, Ryuhei So, Masahiro Banno, Junji Kumasawa, Hidehiro Someko, Shunsuke Taito, Teruhiko Terasawa, Yasushi Tsujimoto, Yusuke Tsutsumi, Yoshitaka Wada, Toshi A. Furukawa

**Author notes:** **Corresponding author:** Toshi A. Furukawa Department of Health Promotion and Human Behavior, Kyoto University Graduate School of Medicine/School of Public Health, Kyoto, Japan. **Author Contributions** YK had full access to all the data in the study and took responsibility for the integrity of the data and the accuracy of the data analysis. Study concept and design: YK, RS, MB, JK, ST, TT, YT, YT, YW, and TAF. Acquisition of data: YK. Drafting of the manuscript: YK. All authors gave final approval of the version to be published and agreed to be accountable for all aspects of this work. **Data Availability Statement** The data that support the findings of this study are openly available at (https://github.com/youkiti/ARE/).

## Abstract

Systematic reviews (SRs) are a critical component of evidence-based medicine, but the process of screening titles and abstracts is time-consuming. This study aimed to develop and externally validate a method using large language models to classify abstracts for diagnostic test accuracy (DTA) systematic reviews, thereby reducing the human workload. We used a previously collected dataset for developing DTA abstract classifiers and applied prompt engineering. We developed an optimized meta-prompt for Generative Pre-trained Transformer (GPT)-3.5-turbo and GPT-4 to classify abstracts. In the external validation dataset 1, the prompt with GPT-3.5 turbo showed a sensitivity of 0.988, and a specificity of 0.298. GPT-4 showed a sensitivity of 0.982, and a specificity of 0.677. In the external validation dataset 2, GPT-3.5 turbo showed a sensitivity of 0.919, and a specificity of 0.434. GPT-4 showed a sensitivity of 0.806, and a specificity of 0.740. If we included eligible studies from among the references of the identified studies, GPT-3.5 turbo had no critical misses, while GPT-4 had some misses. Our study indicates that GPT-3.5 turbo can be effectively used to classify abstracts for DTA systematic reviews. Further studies using other dataset are warranted to confirm our results. Additionally, we encourage the use of our framework and publicly available dataset for further exploration of more effective classifiers using other LLMs and prompts (https://github.com/youkiti/ARE/).

**Hightlights:** *What is already known:*

- Title and abstract screening in systematic reviews (SRs) consumes significant time.
- Several attempts using machine learning to reduce this process in diagnostic test accuracy (DTA) SRs exist, but they have not yielded positive results in external validation.

*What is new:*

- We aimed to develop and externally validate optimized meta-prompt for GPT-3.5-turbo and GPT-4 to classify abstracts for DTA SRs.
- Through an iterative approach across three training datasets, an optimal meta-prompt capable of identifying DTA studies with remarkable sensitivity and specificity was developed.
- The accuracy reproduced in the external validation datasets.

*Potential Impact for Readers:*

- The developed meta-prompt can lessen the need for humans to read abstracts for DTA SRs, saving significant time and resources.

## 1. Introduction

Title and abstract screening in systematic reviews (SRs) requires much time and efforts. Several attempts using machine learning to facilitate this process exist (1,2). Some machine learning models succeeded in intervention and update SRs, but no cases in diagnostic test accuracy (DTA) SRs. In our own previous study, we used the Bidirectional Encoder Representations from Transformers (BERT), which was released in 2018 (3), to develop a model to classify abstracts in DTA SRs. The results were unsatisfactory in the external validation (4).

The launch of Chat Generative Pre-trained Transformer (ChatGPT) in November 2022 has boosted the already high interest in large language models (LLMs) (5). LLMs are machine learning models specifically trained on text data to process and generate human-like text (6). When applying LLMs, there are two techniques: fine tuning and prompt engineering (7). Fine tuning involves training an existing LLM on a new dataset to improve it for a specific task. While this technique is less expensive than creating a new LLM from scratch, it still requires significant time and computational resources. Therefore, more research efforts have been expended on prompt engineering (8–10). Prompt engineering allows for better results from a LLM without additional training by adding what is known as a meta-prompt—a task-specific instruction—in the input. We are aware of one application of prompt engineering to screen references for intervention reviews (11) so far.

However, the accuracy of LLM as a DTA abstract classifier remains uncertain. Our study aimed to develop and externally validate optimized meta-prompts for GPT-3.5-turbo and GPT-4 to classify abstracts for DTA SRs. GPT-3.5-turbo is a version of the GPT model developed by OpenAI. It powers the freely accessible ChatGPT. GPT-4 follows GPT-3.5-turbo as a more advanced model.

## 2. Methods

### 2.1 Preparation of datasets

We used the previously collected dataset for developing the DTA abstract classifier (11). We defined a DTA study as an original study that evaluated a test against a clinical reference standard for humans (13). We classified multivariable diagnostic prediction model studies as DTA studies, but prognostic prediction model studies, that measured predictors and outcomes at different time points as non-DTA studies (14). We classified modeling studies, studies that assessed diagnostic training for medical professionals, and case series (e.g., studies without controls, such as following polymerase chain reaction results of specific patients) as non-DTA studies.

We retrieved various DTA systematic reviews (SRs) from the EPPI-Centre COVID-19: a living systematic map of the evidence (12). These systematic reviews addressed malignancy, gastrointestinal disorders, respiratory disorders, emergency care, neurology, and infectious disease.

The dataset consisted of Microsoft Excel files, including serial numbers, titles, abstracts, and binary reference labels of true (DTA) and false (non-DTA) values. As the reference standard, we used the abstract lists that required manual full-text review when the original DTA SR was conducted. As an additional analysis, we used the included articles after the full-text review as the reference standard in the external validation dataset 2. We used titles and abstracts as predictors.

From 67,979 abstracts used in our previous study (4), which contained 1,575 DTA study abstracts, we conducted stratified sampling for the train dataset 1 (n = 100, 25 DTA abstracts, and 75 non-DTA abstracts). (Figure 1) In addition, we randomly sampled the train dataset 2 (n = 500), and the train dataset 3 (n = 1,000) from among the 1575 DTA studies. These three datasets were used for the development of a meta-prompt to select DTA abstracts. We limited the number of abstracts in the train datasets to decrease data processing time and cost. For external validation of the meta-prompt, we used the same dataset including 7,721 abstracts, including 166 DTA abstracts as used in the previous study (external validation dataset 1) (15). In addition, we used another dataset including 1023 abstracts and 124 DTA abstracts from a DTA SR (external validation dataset 2) (16).

**Figure 1.**
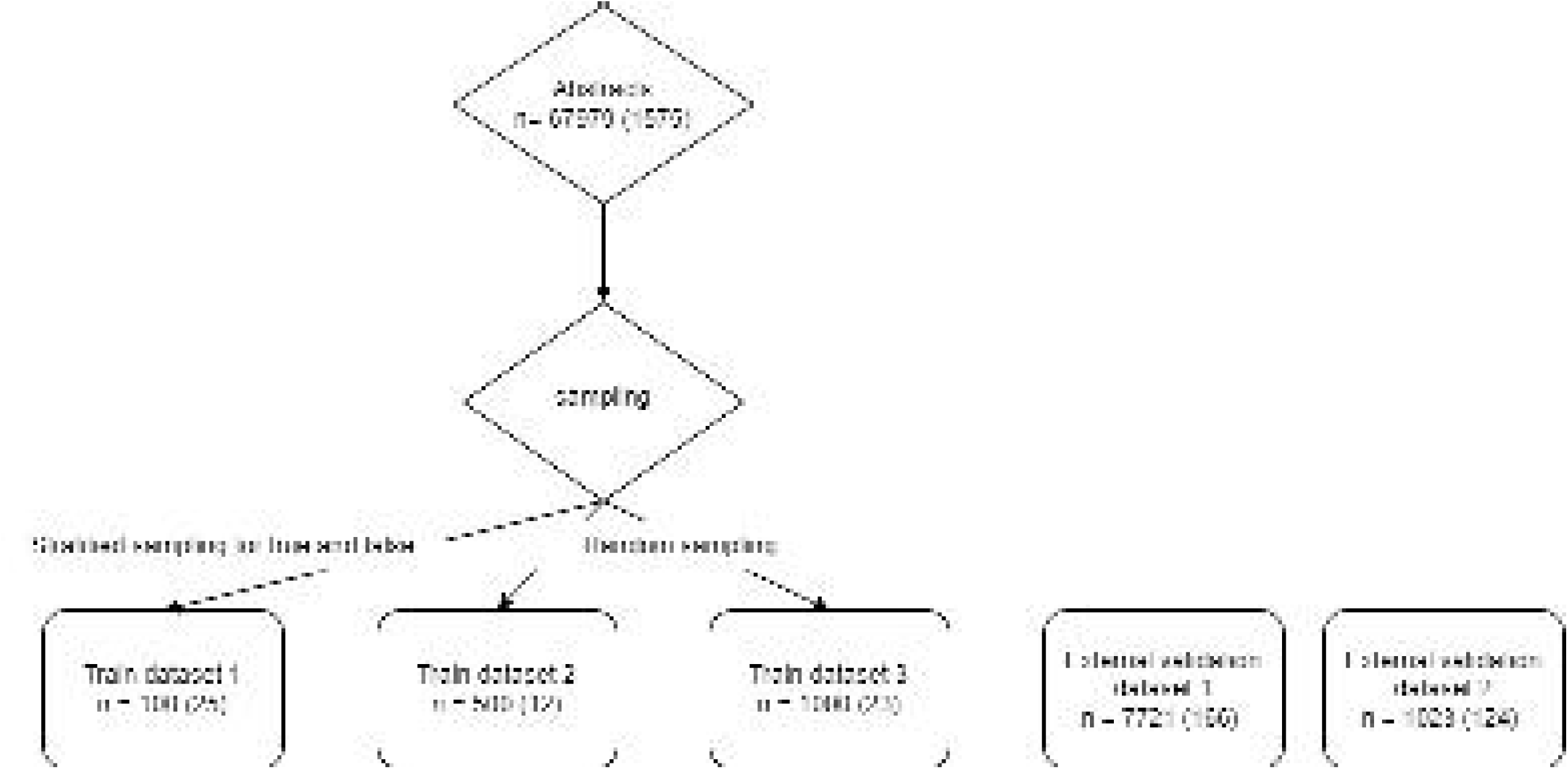
Preparation of datasets. n = number of abstracts (number of diagnostic test accuracy abstracts)

### 2.2 Overview of four-step approach for abstract screening enhancement

In this study, we undertook a four-step approach for abstract review enhancement of diagnostic test accuracy (DTA) abstracts. First, we began by developing a meta-prompt using the Azure OpenAI application programming interface (API), optimizing it for accurate labeling of DTA abstracts (17). Second, we explored the optimal temperature setting for the meta-prompt to achieve the desired outputs. The temperature is the parameter that controls the randomness of the GPT (15). Third, we conducted an external validation using two datasets. Fourthly, we assessed the reproducibility of the model’s outputs and iterative accuracy enhancement (Figure 2).

**Figure 2.**
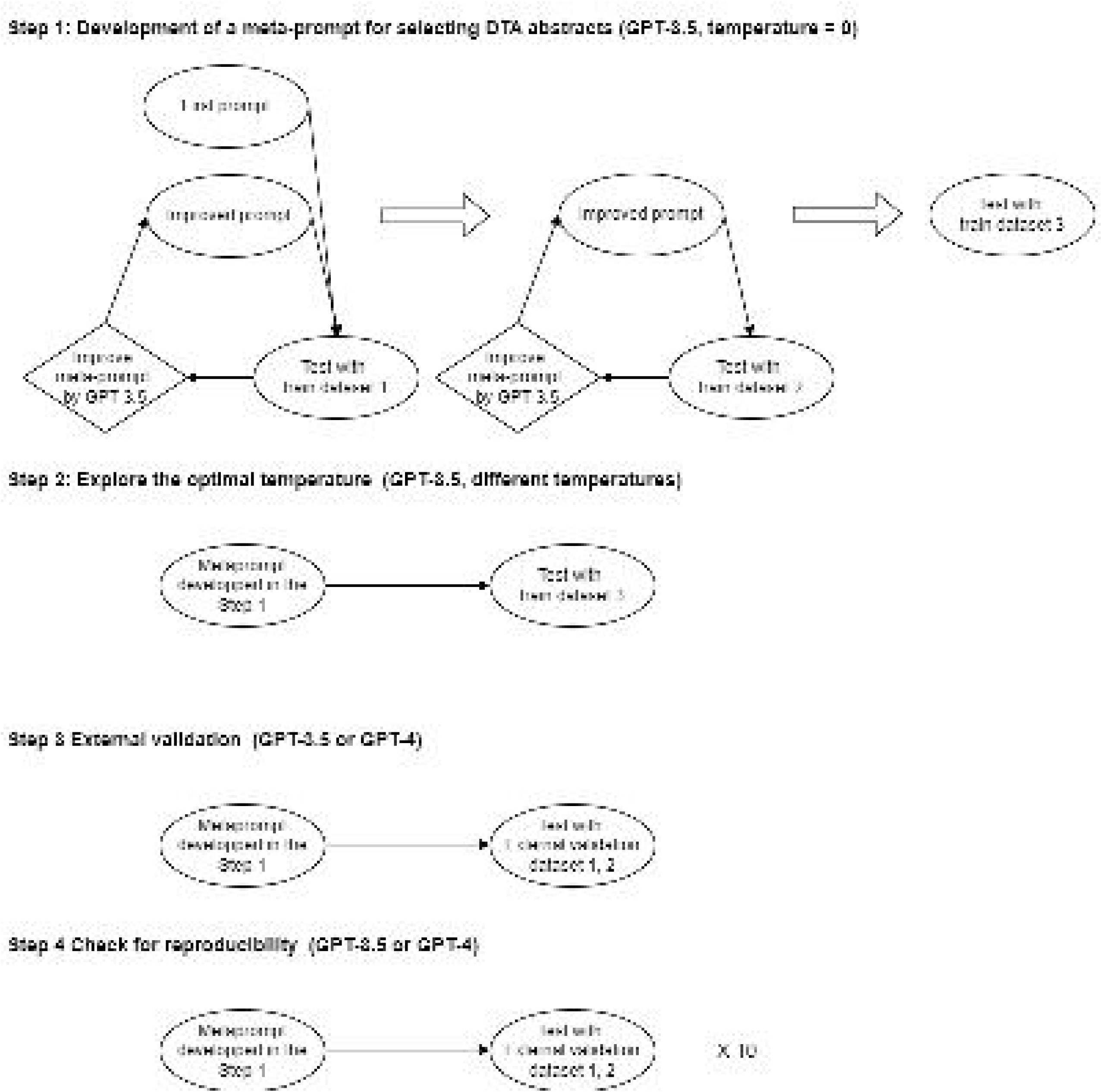
Each step to develop, externally validate, and checking for reproducibility of the meta-prompts. GPT: Generative Pre-trained Transformer

### 2.3 Step 1: Development of a meta-prompt for selecting DTA abstracts

We used Azure OpenAI API which provides access to GPT-3.5 turbo and GPT-4. The input included a meta-prompt, a title and an abstract, and the temperature parameter. The meta-prompt was to label whether the inputted abstracts were DTA abstracts or not. One title-and-abstract was retrieved from each line of dataset. The temperature is the parameter that controls the randomness of the GPT (18). The temperature has a valid range from 0.0 inclusive to 2.0 exclusive. Higher values will make output more random while lower values will make results more focused and deterministic. We set the temperature as 0 for the accurate labeling. The output was a label of true or false. We used GPT-3.5 turbo to develop a meta-prompt (Figure 2 and 3).

**Figure 3.**
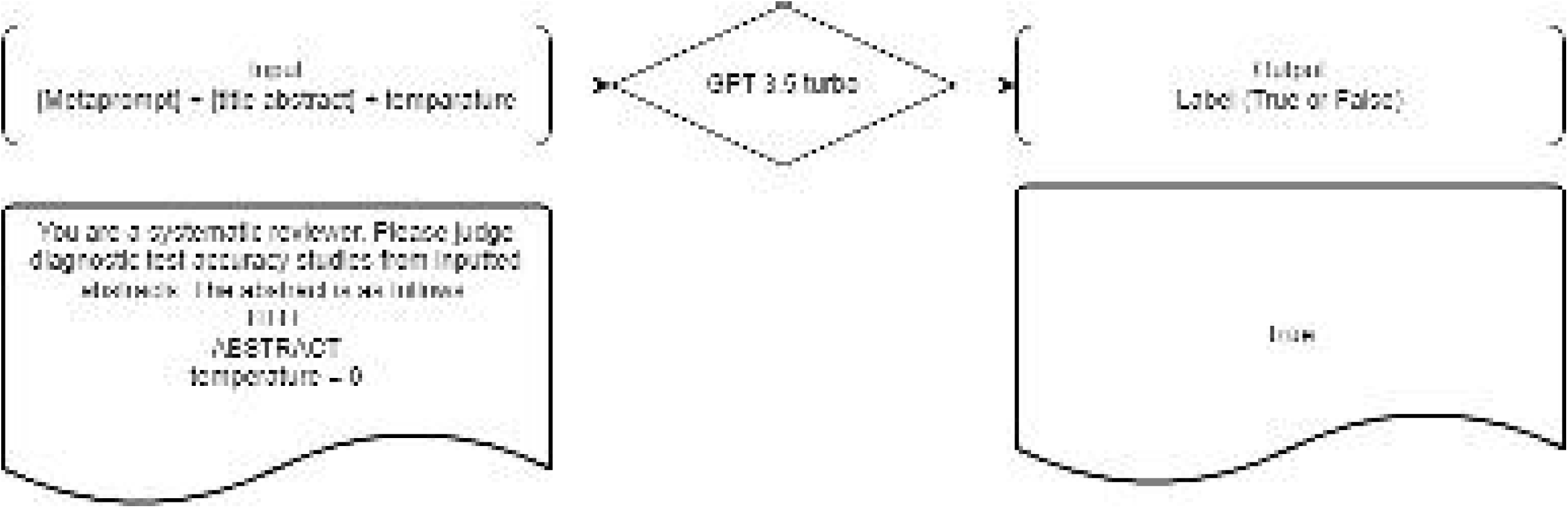
Schema of input and output for large language models. GPT: Generative Pre-trained Transformer The square below shows an example input and output.

From the predicted label, we calculated the sensitivity and the specificity and the proportion of error as performance measures. Then we asked the GPT-3.5 turbo to improve the meta-prompt (Figure 2). The improvement meta-prompt was as follows:

> Please become my prompt engineer. Your goal is to help me create the best prompts for systematic review of diagnostic test accuracy. The prompts will be used by you, ChatGPT. Please rewrite inputted meta-prompt to achieve sensitivity > 0.9 and specificity > 0.4 and error proportion < 0.1.

We selected the above cutoffs based on a previous study that investigated the search filters for systematic reviews (19). The error meant that when a response other than true or false occurs three times in an abstract, which included communication errors.

Firstly, we ran the experiment 10 times with the train dataset 1 and chose the best one. Secondly, we ran the experiment 10 times with the train dataset 2 and chose the best one. Thirdly, we tested with the train dataset 3.

### 2.4 Step 2: Explore the optimal temperature

As mentioned above, the temperature was set to 0 in the Step 1. To explore the optimal temperature, we used the optimal meta-prompt and changed temperature as 0,0.4,0.8, 1.2, and 1.6. We evaluated the results with sensitivity, specificity, and error proportion (Figure 2).

### 2.5 Step 3: External validation

For the external validation, we used the optimal meta-prompt developed in the step 1 with the external validation dataset 1 and 2. We used GPT-3.5 turbo and GPT-4. We evaluated the results with sensitivity, specificity, error proportion, and number needed to screen (Figure 2). Number needed to screen is the number to identify 1 reference to undergo full-text screening during title and abstract screening (20). For the external validation dataset 2, we assessed the accuracy using abstracts deemed ‘true’ after a full-text review by human experts as the reference standard (RS2). Additionally, we examined the characteristics of abstracts that turned out to be false negatives for the RS2.

### 2.6 Step 4: Check for reproducibility

Large language models like GPT have inherent non-determinism (21,22). Outputs remain non-deterministic, even at a temperature of 0 (23). Hence, we checked the reproducibility of GPT-3.5 turbo and GPT-4 using the same meta-prompt ten times for the external validation dataset 1 and 2 (Figure 2).

For the external validation dataset 2, we evaluated the performance enhancement when combining results by considering an abstract as ‘true’ if it was deemed ‘true’ in at least one of the ten trials.

### 2.7 Development environment

We used Google Collaboratory, a Python-based data analysis and machine learning tool that can be executed in a web browser (24). We used the Azure OpenAI API version "2023-07-01-preview". We used "gpt-35-turbo-0613" as GPT-3.5 turbo, "gpt4-0613" as GPT-4. Our code and datasets are made available at GitHub (https://github.com/youkiti/ARE/).

## 3. Results

### 3.1 Step 1: Development of a meta-prompt for selecting DTA abstracts

We developed the first meta-prompt and improved the meta-prompt ten times with the training dataset 1 (n = 100). Then selected the #8 prompt based on the balance of sensitivity and specificity (Table 1, Supplemental table 1).

**Table 1.**
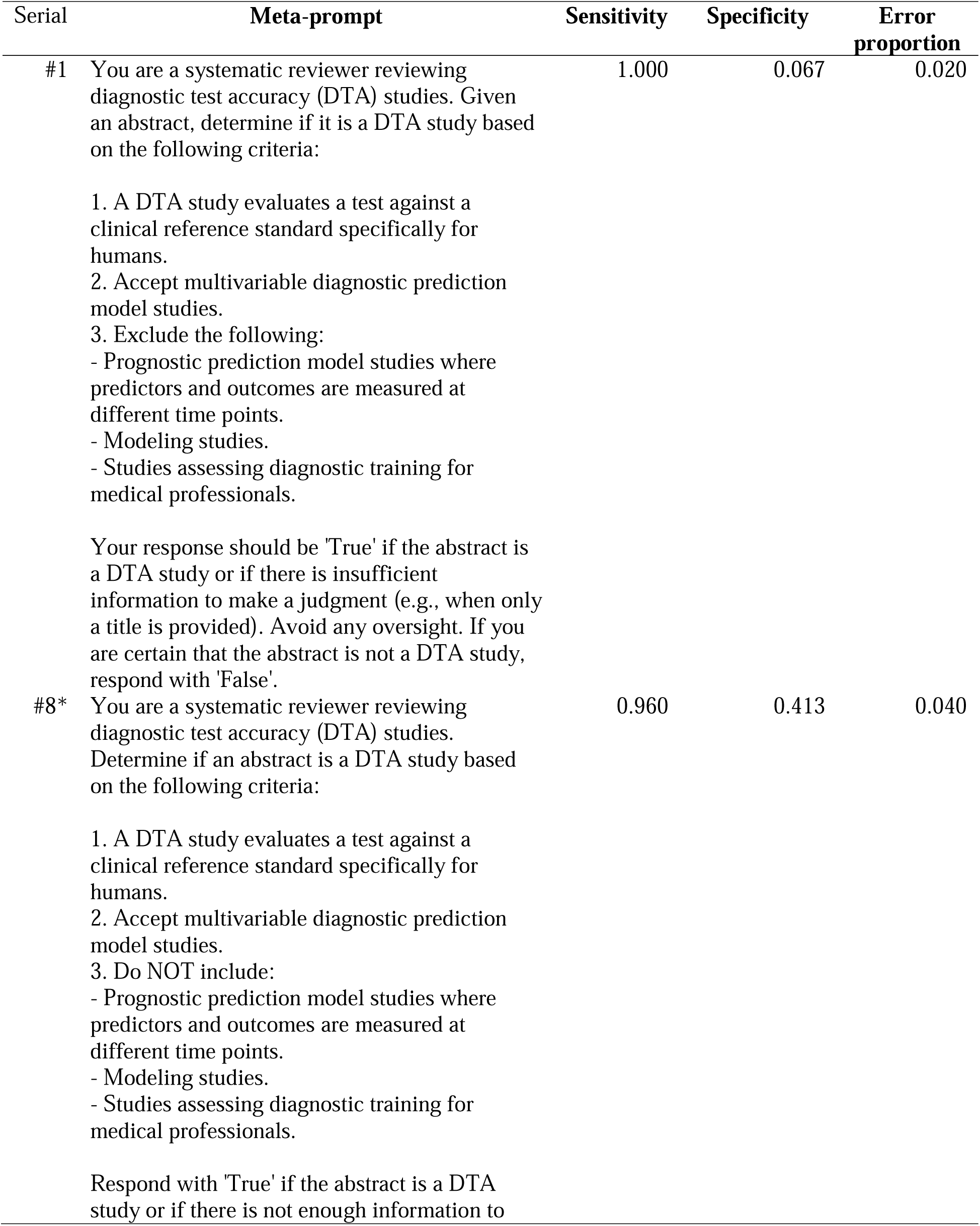
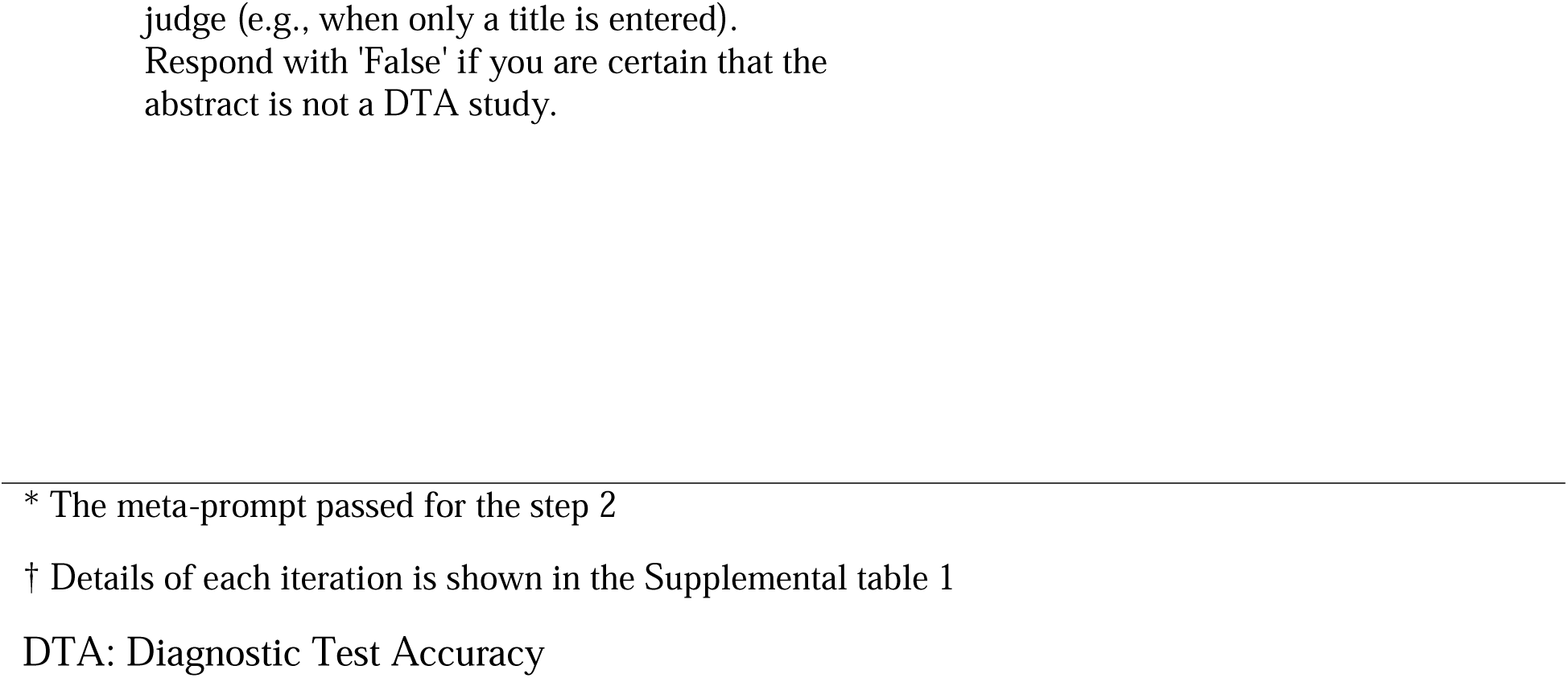
Accuracy of meta-prompts for selecting DTA abstracts in the training dataset 1.

Using the #8 meta-prompt, we improved the meta-prompt with the training dataset 2 (n = 500) (Table 2). The #3 prompt achieved a sensitivity of 0.917, a specificity of 0.527, and an error proportion of 0.010.

**Table 2.**
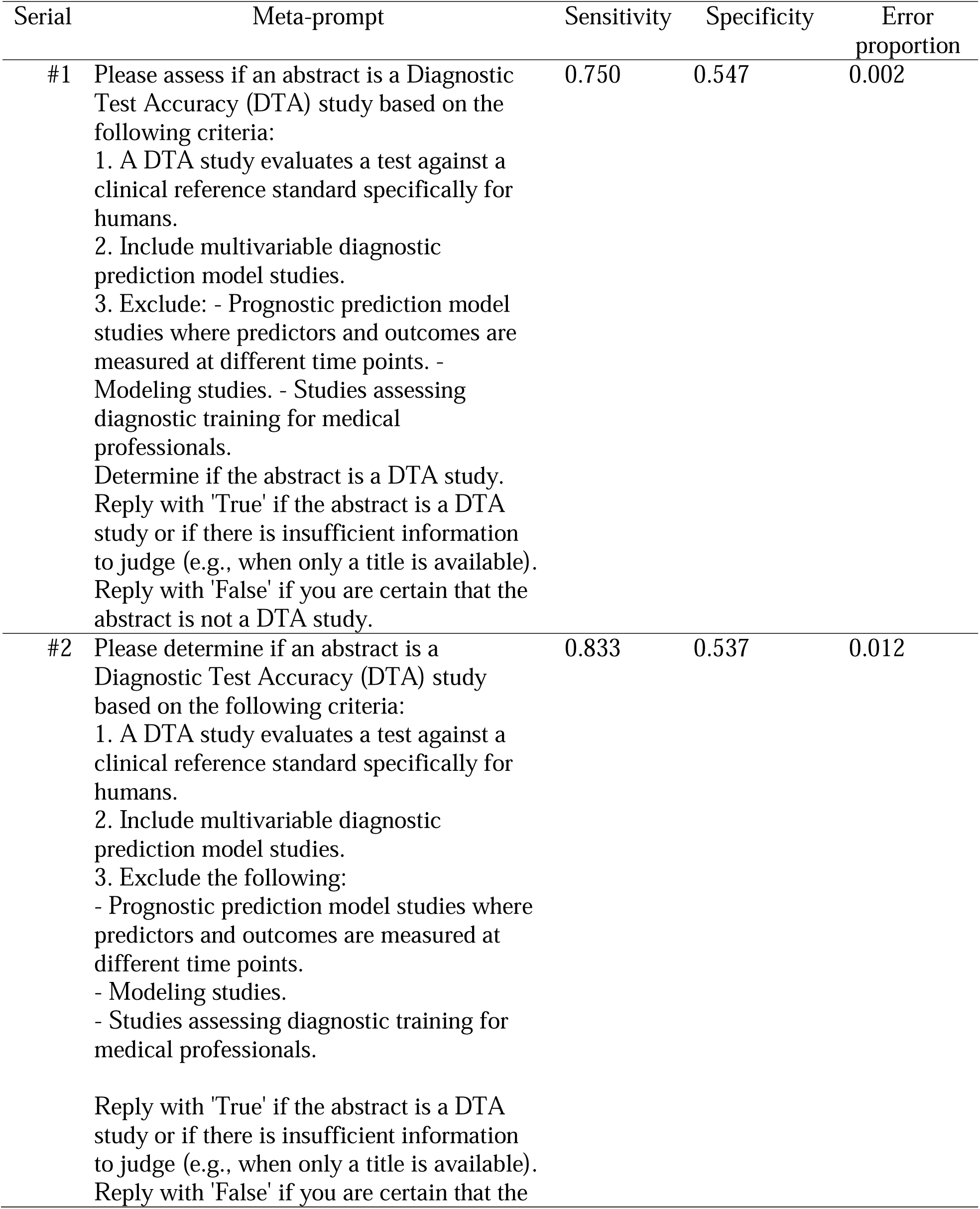
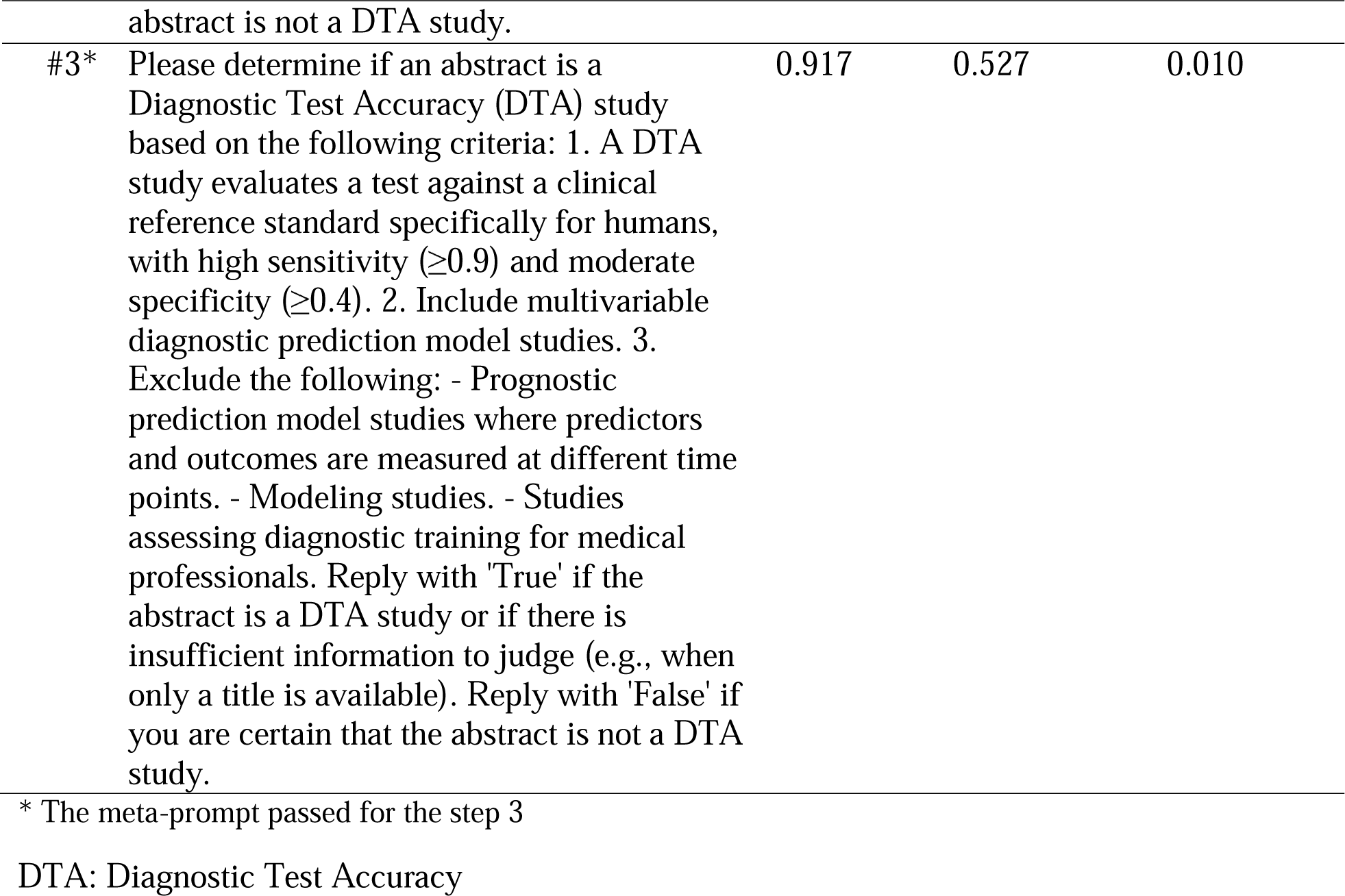
Accuracy of meta-prompts for selecting DTA abstracts in the training dataset 2.

| Serial | Meta-prompt | Sensitivity | Specificity | Error proportion |
| --- | --- | --- | --- | --- |
| #1 | Please assess if an abstract is a Diagnostic Test Accuracy (DTA) study based on the following criteria:<br>1. A DTA study evaluates a test against a clinical reference standard specifically for humans.<br>2. Include multivariable diagnostic prediction model studies.<br>3. Exclude: - Prognostic prediction model studies where predictors and outcomes are measured at different time points. - Modeling studies. - Studies assessing diagnostic training for medical professionals.<br>Determine if the abstract is a DTA study. Reply with 'True' if the abstract is a DTA study or if there is insufficient information to judge (e.g., when only a title is available). Reply with 'False' if you are certain that the abstract is not a DTA study. | 0.750 | 0.547 | 0.002 |
| #2 | Please determine if an abstract is a Diagnostic Test Accuracy (DTA) study based on the following criteria:<br>1. A DTA study evaluates a test against a clinical reference standard specifically for humans.<br>2. Include multivariable diagnostic prediction model studies.<br>3. Exclude the following:<br>- Prognostic prediction model studies where predictors and outcomes are measured at different time points.<br>- Modeling studies.<br>- Studies assessing diagnostic training for medical professionals.<br>Reply with 'True' if the abstract is a DTA study or if there is insufficient information to judge (e.g., when only a title is available). Reply with 'False' if you are certain that the<br>abstract is not a DTA study. | 0.833 | 0.537 | 0.012 |
| #3* | Please determine if an abstract is a Diagnostic Test Accuracy (DTA) study based on the following criteria: 1. A DTA study evaluates a test against a clinical reference standard specifically for humans, with high sensitivity ($\geq 0.9$) and moderate specificity ($\geq 0.4$). 2. Include multivariable diagnostic prediction model studies. 3. Exclude the following: - Prognostic prediction model studies where predictors and outcomes are measured at different time points. - Modeling studies. - Studies assessing diagnostic training for medical professionals. Reply with 'True' if the abstract is a DTA study or if there is insufficient information to judge (e.g., when only a title is available). Reply with 'False' if you are certain that the abstract is not a DTA study. | 0.917 | 0.527 | 0.010 |

\* The meta-prompt passed for the step 3
DTA: Diagnostic Test Accuracy

We tested the #3 meta-prompt with the training dataset 3 (n = 1000). The prompt achieved a sensitivity of 0.913, a specificity of 0.416, and an error proportion of 0.000. To enhance the prompt, we omitted the numbers for the thresholds of sensitivity and specificity. The final meta-prompt was as follows:

> Please determine if an abstract is a Diagnostic Test Accuracy (DTA) study based on the following criteria:
>
> 1. A DTA study evaluates a test against a clinical reference standard specifically for humans, with very high sensitivity and reasonable specificity.
> 2. Include multivariable diagnostic prediction model studies.
> 3. Exclude the following:
>   - Prognostic prediction model studies where predictors and outcomes are measured at different time points.
>   - Modeling studies.
>   - Studies assessing diagnostic training for medical professionals.
>
> Reply with ‘True’ if the abstract is a DTA study or if there is insufficient information to judge (e.g., when only a title is available). Reply with ‘False’ if you are certain that the abstract is not a DTA study.

In the training dataset 3, the prompt achieved a sensitivity of 0.938, a specificity of 0.514, and an error proportion of 0.010 (Table 3).

**Table 3.** Accuracy of the final meta-prompt at different temperatures with GPT-3.5 turbo in the training dataset 3.

| Temperature | Sensitivity | Specificity | Error proportion |
| --- | --- | --- | --- |
| 0 | 0.938 | 0.514 | 0.010 |
| 0.4 | 0.875 | 0.518 | 0.006 |
| 0.8 | 0.813 | 0.491 | 0.009 |
| 1.2 | 0.813 | 0.480 | 0.025 |
| 1.6 | 0.688 | 0.449 | 0.109 |

GPT: Generative Pre-trained Transformer

### 3.2 Step 2: Explore the optimal temperature

We observed a decrease in sensitivity as the temperature increased (Table 3).

### 3.3 Step 3: External validation

For the final meta-prompt tested on the external validation dataset 1, GPT-3.5 turbo showed a sensitivity of 0.988, a specificity of 0.298, and an error rate of 0.008, while GPT-4 showed a sensitivity of 0.982, a specificity of 0.677, and an error proportion of 0.008. For the final meta-prompt tested on the external validation dataset 2, GPT-3.5 turbo showed a sensitivity of 0.919, a specificity of 0.434, and an error proportion of 0.005, while GPT-4 showed a sensitivity of 0.806, a specificity of 0.740, and an error proportion of 0.008 (Table 4).

**Table 4.** Reproducibility test from 10 experiments for the external validation dataset 1.

| **Serial** | **model** | **Sensitivity** | **Specificity** | **Error proportion** |
| --- | --- | --- | --- | --- |
| #1 | GPT-3.5 | 0.988 | 0.298 | 0.008 |
| #2 | GPT-3.5 | 0.988 | 0.297 | 0.005 |
| #3 | GPT-3.5 | 0.994 | 0.286 | 0.008 |
| #4 | GPT-3.5 | 0.982 | 0.288 | 0.009 |
| #5 | GPT-3.5 | 0.988 | 0.288 | 0.009 |
| #6 | GPT-3.5 | 0.988 | 0.292 | 0.011 |
| #7 | GPT-3.5 | 0.994 | 0.289 | 0.011 |
| #8 | GPT-3.5 | 0.988 | 0.292 | 0.010 |
| #9 | GPT-3.5 | 0.988 | 0.294 | 0.010 |
| #10 | GPT-3.5 | 0.994 | 0.295 | 0.010 |
| #1 | GPT-4 | 0.982 | 0.677 | 0.000 |
| #2 | GPT-4 | 0.976 | 0.678 | 0.002 |
| #3 | GPT-4 | 0.976 | 0.677 | 0.002 |
| #4 | GPT-4 | 0.976 | 0.678 | 0.002 |
| #5 | GPT-4 | 0.982 | 0.677 | 0.001 |
| #6 | GPT-4 | 0.988 | 0.679 | 0.000 |
| #7 | GPT-4 | 0.994 | 0.679 | 0.000 |
| #8 | GPT-4 | 0.994 | 0.678 | 0.000 |
| #9 | GPT-4 | 0.982 | 0.678 | 0.000 |
| #10 | GPT-4 | 0.988 | 0.681 | 0.000 |

GPT: Generative Pre-trained Transformer

On the external validation dataset 1, the baseline number needed to screen was 46.5. The number reduced to 33.3 with GPT-3.5 turbo, and to 16.0 with GPT-4. In external validation dataset 2, the number needed to screen reduced from 8.25 to 5.45 with GPT-3.5 turbo, and to 3.34 with GPT-4.

When we used the included articles after the full-text review as the reference standard (RS2), in the external validation dataset 2, GPT-3.5 turbo showed a sensitivity of 0.963 and a specificity of 0.406, while GPT-4 showed a sensitivity of 0.889 and a specificity of 0.689. In other words, GPT-3.5 missed one abstract from 27 abstracts included in the reviews and GPT-4 missed three abstracts. The one abstract that GPT-3.5 turbo missed (25) was referenced in another included article (26). Two of three abstracts that GPT-4 missed were not detectable by citation search of included articles (27,28).

### 3.4 Step 4: Check for reproducibility

We observed no remarkable differences in sensitivity, specificity, and error proportion between GPT-4 and GPT-3.5 in both external validation datasets during the ten experiments (Table 4 and 5).

**Table 5.**
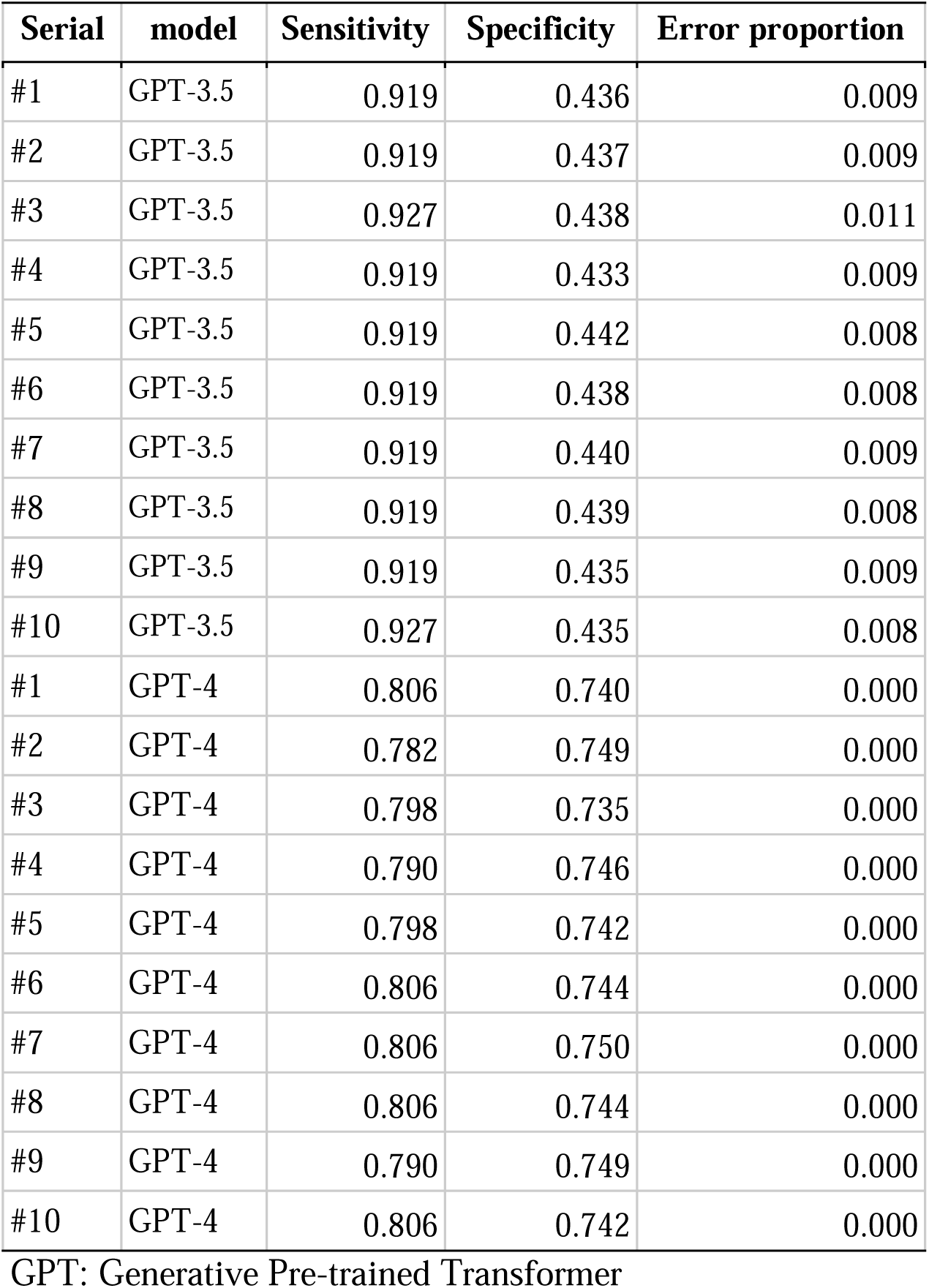
Reproducibility test from 10 experiments for the external validation dataset 2.

| **Serial** | **model** | **Sensitivity** | **Specificity** | **Error proportion** |
| --- | --- | --- | --- | --- |
| #1 | GPT-3.5 | 0.919 | 0.436 | 0.009 |
| #2 | GPT-3.5 | 0.919 | 0.437 | 0.009 |
| #3 | GPT-3.5 | 0.927 | 0.438 | 0.011 |
| #4 | GPT-3.5 | 0.919 | 0.433 | 0.009 |
| #5 | GPT-3.5 | 0.919 | 0.442 | 0.008 |
| #6 | GPT-3.5 | 0.919 | 0.438 | 0.008 |
| #7 | GPT-3.5 | 0.919 | 0.440 | 0.009 |
| #8 | GPT-3.5 | 0.919 | 0.439 | 0.008 |
| #9 | GPT-3.5 | 0.919 | 0.435 | 0.009 |
| #10 | GPT-3.5 | 0.927 | 0.435 | 0.008 |
| #1 | GPT-4 | 0.806 | 0.740 | 0.000 |
| #2 | GPT-4 | 0.782 | 0.749 | 0.000 |
| #3 | GPT-4 | 0.798 | 0.735 | 0.000 |
| #4 | GPT-4 | 0.790 | 0.746 | 0.000 |
| #5 | GPT-4 | 0.798 | 0.742 | 0.000 |
| #6 | GPT-4 | 0.806 | 0.744 | 0.000 |
| #7 | GPT-4 | 0.806 | 0.750 | 0.000 |
| #8 | GPT-4 | 0.806 | 0.744 | 0.000 |
| #9 | GPT-4 | 0.790 | 0.749 | 0.000 |
| #10 | GPT-4 | 0.806 | 0.742 | 0.000 |

GPT: Generative Pre-trained Transformer

As a result of combining multiple evaluations for one abstract, we observed the minimal improvement in the external validation dataset 2 using GPT-3.5 turbo and GPT-4 (Table 6).

**Table 6.** Accuracy of the final meta-prompt when combining the results with GPT-3.5 turbo and GPT-4 in the external validation dataset 2.

| Combined experiments<br>count | GPT-3.5 turbo |  | GPT-4 |  |
| --- | --- | --- | --- | --- |
|  | Sensitivity | Specificity | Sensitivity | Specificity |
| 1 | 0.919 | 0.446 | 0.806 | 0.740 |
| 2 | 0.919 | 0.439 | 0.815 | 0.734 |
| 3 | 0.927 | 0.437 | 0.815 | 0.722 |
| 4 | 0.927 | 0.434 | 0.815 | 0.721 |
| 5 | 0.927 | 0.430 | 0.815 | 0.720 |
| 6 | 0.927 | 0.427 | 0.815 | 0.719 |
| 7 | 0.927 | 0.427 | 0.823 | 0.717 |
| 8 | 0.927 | 0.426 | 0.823 | 0.716 |
| 9 | 0.927 | 0.420 | 0.823 | 0.714 |
| 10 | 0.935 | 0.419 | 0.823 | 0.714 |

GPT: Generative Pre-trained Transformer

## 4. Discussion

We developed and externally validated the meta-prompt to classify abstracts for new DTA systematic reviews. Through an iterative approach across three training datasets, we developed an optimal meta-prompt capable of identifying DTA studies with remarkable sensitivity and specificity. The temperature parameter, when set to 0, demonstrated the best performance. In the external validation dataset 1, using the same meta-prompt, GPT-3.5 turbo and GPT-4 showed almost the same sensitivity and error proportion. In the external validation dataset 2, GPT-3.5 and GPT-4 showed worse sensitivity. However, combining citation search, GPT-3.5 turbo had no substantive misses. GPT-4 had some misses. As a result of the check for reproducibility, we observed no remarkable differences in results across the 10 serial experiments. Combining multiple evaluations for one abstract did not notably improve performance.

Our results are better than in our previous study that used machine learning. In our research using the fine-tuned model of BERT, the sensitivity in the external validation set was less than 0.4 (26). In this study, both GPT-3.5 turbo and GPT-4 achieved a sensitivity exceeding 0.96. The performance is equivalent to existing RCT search filters (19). GPT-3.5 turbo had similar or better sensitivity, while GPT-4 demonstrated better specificity. Regarding time and cost, as of October 2023, using GPT-3.5 turbo API on the fastest setting of S0 Standard Azure took 1 min 18 seconds to process 100 abstracts and 0.09 dollars cost. In contrast, GPT-4 API took 2 min 44 seconds and 1.7 dollars cost.

Our results indicate a lack of strict reproducibility in outputs of LLMs, even with zero temperature settings in binary labeling tasks. In other words, occasionally, LLMs produce different outputs from the same input. However, the lack of reproducibility is the same even when a human makes the abstract review. In fact, if the same person classifies abstracts one week later, the results do not necessarily match (29). Drawing parallels to epidemiological studies, it’s worth noting that LLMs inherently involve measurement errors (30). The precise nature of the inconsistency of LLMs, be it systematic or random, remains unclear. To effectively assess the performance of LLMs, understanding the non-deterministic nature will help address this issue. Reflecting on our research objectives, the variability in judgment did not substantially affect the sensitivity.

We position our study as a type of prompt engineering by the LLM itself. Researchers are exploring a framework to enhance meta-prompts by presenting specific tasks and meta-prompts and their scores to the LLM. Researchers have applied this framework to mathematical problems (31) and simple natural language processing tasks (32,33). In systematic reviews, the potential exists to implement appropriate prompt engineering using LLM for tasks where the dataset can provide correct answers.

Our study has several limitations. Firstly, we have yet to validate our findings on other datasets. Future studies are warranted to test our results on alternative datasets to ascertain the generalizability of our conclusions. Secondly, there remains an unanswered question regarding the efficacy of the meta-prompts in relation to other LLMs. The best meta-prompts might be different for each LLM (34). "Closed” OpenAI LLMs can only be accessed through the API and cannot be downloaded to run on a researchers’ computer. Therefore, there is a risk they may change or even become inaccessible in the future. Researchers should have alternative open LLMs that can be run on their own server. Lastly, our current study has scoped its focus predominantly on the study design. For abstract review enhancement, further studies are warranted to determine if a meta-prompt considering other DTA study elements, such as participants and index tests, can reduce the number needed to screen in new reviews (11).

## 5. Conclusions

In conclusion, we developed and externally validated the meta-prompt to reduce the burden for humans to read abstracts when conducting DTA SRs. Considering situations where cost and sensitivity are prioritized, we recommend systematic reviewers to use GPT-3.5 turbo and our meta-prompt for title and abstract screening of DTA reviews. Further studies using other dataset are warranted to confirm our results.

## Data Availability

All data produced are available online at (https://github.com/youkiti/ARE/)

## Acknowledgment

The authors underwent editing using GPT-0614. All authors reviewed and edited the final manuscript. The responsibility for the content of this article rests solely with the authors.

